# A New Hematological Prognostic Index For Covid-19 Severity

**DOI:** 10.1101/2021.02.11.21251285

**Authors:** Abdulmajeed A. Hammadi, Adnan M. Al Jubouri, Ghassan Ahmed, Ali H. Hayyawi, Khalil Kareem, Faiq I. Gorial, Muhammed Waheeb Salman, Mohammed Ghanim, Basil F. Jameel, Ali M. Jawad, Hassan M. Abbas, Ali A. Al-Gharawi, Jawad I Al-shareef, Chasib L. Ali, Kawthar F. Nasser, Mohammad Y. Abdulrazaq

## Abstract

**Background:** severe acute respiratory syndrome coronavirus 2 (SARS-CoV-2) or Covid-19 is a nationwide public health emergency with significant impact on human life.

**Objective:** To develop a new simple hematological prognostic index for Covid-19 severity state.

**Patients and methods:** This observational cross sectional study was conducted on 250 patients with Covid-19 disease. Age, gender, and severity of Covid −19 were recorded. Complete blood count and lactate dehydrogenase were measured.

New index: COVID-19 severity-Iraqi-index = CSI index to predict COVID-19 severity.

CSI index is monocyte/lymphocyte absolute counts multiplied by LDH (lactate dehydrogenase)/upper normal reference laboratory range of LDH value.

**Results:** Mean age of patients was 50.4 + 15.1 years. Majority of patients were Males 148 (59.2%)..Most of patients were in stage 2 and 3 (> 94%). There was a significant difference between means of White Blood Cells, lymphocytes and Monocytes among the different stages of the disease (P = 0.0001, 0.036, 0.012). There was a significant moderate correlation between the prognostic index and the stage of the disease (r=0.41, p=0.0001).

**Conclusions:** CSI index is a new simple predictor of clinical outcome in patients with covid-19 during early stage of the disease.

## Introduction

Severe acute respiratory syndrome corona virus 2 (SARSCov2) or corona virus 19 COVID 19) pandemic infects more than 50 million people globally with mortality of more than one million all over the world till now. The World Health Organization declared Covid-19 as a pandemic on 11 March 2020. It is reported that around 10 % of the patients progressed toward critical illness (1-3).

Covid −19 is a clinical spectrum of illness ranging from mild form with upper respiratory symptoms including loss of sense of smell or taste to moderate condition with fever and pneumonia to severe lung infiltrations with respiratory failure (4). Many studies pointed to prognostic indices related to variety of factors including: older age, chronic diseases, impaired immunity and many biomarkers like serum ferritin, D dimer, serum lactate dehydrogenase, lymphopenia, albumen level, C reactive protein, procalcitonin and others(5-8).

Since the covid-19 pandemic impacts a pressure on intensive care facilities, it would be important to stratify patients’ admission to those critical care units. (6) It is important to know risk factors and biomarkers that predict disease progression at earlier stages to modify treatment protocols.The aim of this study was to develop a new simple hematological prognostic index for Covid-19 severity state.

## Patients and methods

### Study design

This analytic cross sectional study was conducted at the covid-19 outpatient specialized clinic or admitted to the isolated ward in the hospital in Al-Yarmouk Teaching Hospital and Baghdad Teaching Hospital from June 2020 to November 2020.

The study was approved by the ethical research committee of Department of Medicine, College of Medicine, University of Baghdad with the No. ERC.DoM.CoM,UoB.Ju.2020.1 with date: of June, 2nd, 2020 and was conducted according to the Declaration of Helsinki and its amendments and the Guidelines for Good Clinical Practices issued by the Committee of Propriety Medicinal Product of the European Union. An informed consent was taken from all participants prior to their inclusion in the study.

## Sample selection

### Inclusion criteria

1. Patients with age above 16 years and of any gender
2. Definite diagnosis of COVID-19 according to the WHO classification criteria (18). 18) World Health Organization. Clinical management of COVID-19: interim guidance, 27 May 2020. World Health Organization; 2020
3. Patient understands and agrees to comply with planned study procedures
4. Severity assessment was based on 4 clinical stages according to (COVID-19) Outbreak in China: Summary of a Report of 72_LJ_314 Cases from the Chinese Center for Disease Control and Prevention (1)
  A. Mild Disease: Patients with mild illness may present with symptoms of an upper respiratory tract viral infection. These include dry cough, mild fever, nasal congestion, sore throat, headache, muscle pain, and malaise. It is also characterized by the absence of serious symptoms such as dyspnea. The majority (81%) of COVID-19 cases are mild in severity. Furthermore, radiograph features are also absent in such cases Patients with mild disease can quickly deteriorate into severe or critical cases.
  B. Moderate Disease: These patients present with respiratory symptoms of cough, shortness of breath, and tachypnea. However, no signs and symptoms of severe disease are present.
  C. Severe Disease: Patients with severe disease present with severe pneumonia. Diagnosis is clinical, and complications can be excluded with the help of radiographic studies. Clinical presentations include the presence of severe dyspnea, tachypnea (respiratory rate > 30/minute), respiratory distress, SpO2 _≤_ 93%, PaO2/FiO2 < 300, and/or greater than 50% lung infiltrates within 24 to 48 hours.
  D. Critical disease: In addition, 5% of patients can develop a critical disease with features of respiratory failure, RNAaemia, cardiac injury, septic shock, or multiple organ dysfunctions. Data from the Chinese Centers for Disease Control and Prevention (CDC) suggest that the case fatality rate for critical patients is 49%.

### Exclusion criteria

1. Patients refuse to enrol in the study
2. Pregnancy

### Procedure

A new index: COVID-19 severity-Iraqi-index = CSI index was developed. The CSI index is calculated according to the following formula: Monocyte/lymphocyte absolute counts x LDH (lactate dehydrogenase)/upper normal LDH value This new index is the result of 3 main parameters which are the monocyte, lymphocyte count and lactate dehydrogenase which is the main agreed upon enzyme reflecting the disease severity.

### Statistical issue

data were grouped and analyzed using SPSS v26 program. A comparison of means was performed using ANOVA test for the normally distributed data and Kruskal-Wallis test for variables not follow the normal distribution. A correlation statistic was done to find the correlation coefficient.The median index value for moderate cases was considered as a cutoff value for moderate diseases. The median index value for severe cases was considered as a cutoff value for severe diseases.

## Results

A total of 250 patients with Covid-19 were included in this study. The mean and standard deviation of the age was 50.4 + 15.1 years with range from 16 year to 85 year. Males were more than female 148(59.2%), 102(40.8%) respectively. The majority of patients were in stage 2 and 3 forming together more than 94% of the study group. Stage 1 mild cases were 4 (1, 6%), Stage 2 moderate cases were 112 (44, 8%), stage 3 severe cases were 124 (49, 6%), and stage 4 critical cases were 10 (4%) (Table-1).

**Table 1:** Distribution of the sample according to some variables

| Variables (N=250) |  | Frequency | Percent |
| --- | --- | --- | --- |
| Age group in years |  |  |  |
|  | 16-30 | 35 | 14.0 |
|  | 31-45 | 64 | 25.6 |
|  | 46-60 | 88 | 35.2 |
|  | 61-85 | 63 | 25.2 |
| Gender |  |  |  |
|  | Male | 148 | 59.2 |
|  | Female | 102 | 40.8 |
| Stages |  |  |  |
|  | Stage 1 | 4 | 1.6 |
|  | Stage 2 | 113 | 45.2 |
|  | Stage 3 | 123 | 49.2 |
|  | Stage 4 | 10 | 4.0 |

All patients were improved and discharged from the hospital except four cases were dead.

Measurement of main blood constituents were done for all the patients including automated blood cell counts and a comparison was made in their level in the four stages of the disease. The results shown in table-2 which revealed a significant difference between means of WBC, lymphocytes, monocytes and hemoglobin among the different stages of the disease (P <0.001, p<0.001. p<0.001, p<0.001), but it was not significant for platelets count (P = 0.732) and monocytes (P = 0.065).

Table 3: represented the Spearman’s correlation study. There was a significant correlation between the index and the stage of the disease, but the correlation coefficient was moderate one (0.41), the prediction value was 15%.

**Table 2:**
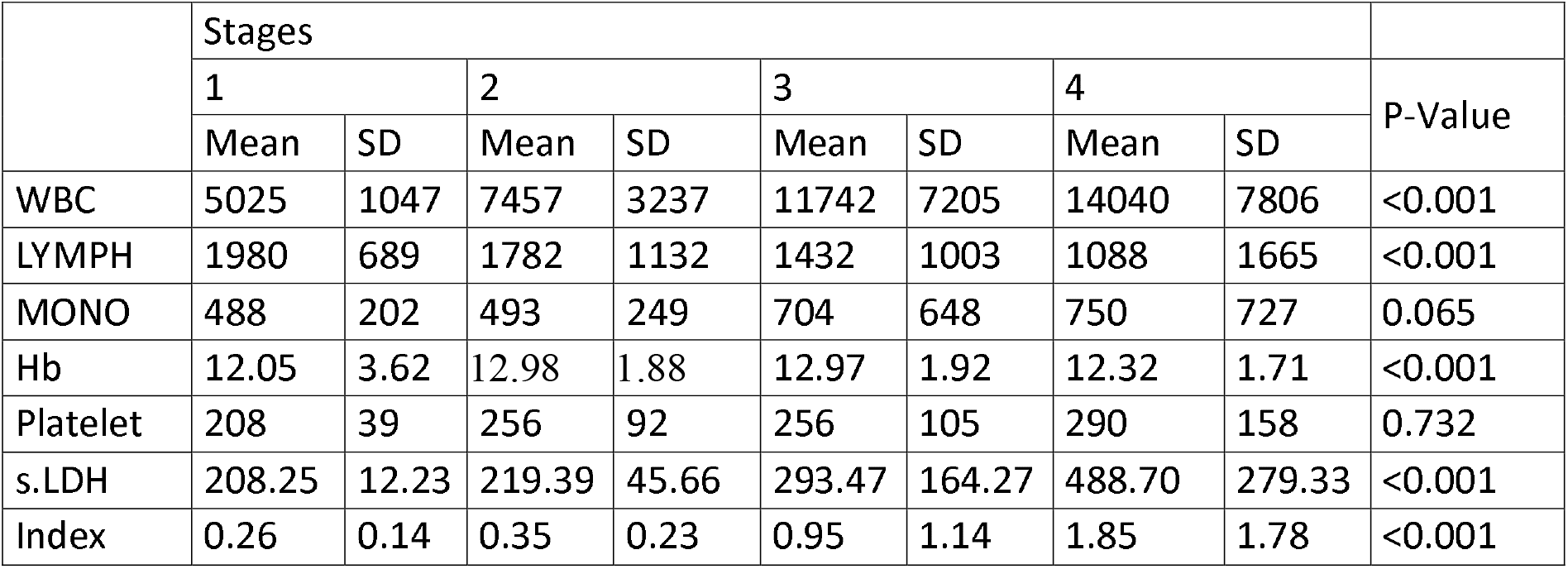
The mean and standard deviation of blood cell counts and serum lactate dehydrogenase in different stages

|  | Stages |  |  |  |  |  |  |  | P-Value |
| --- | --- | --- | --- | --- | --- | --- | --- | --- | --- |
|  | 1 |  | 2 |  | 3 |  | 4 |  |  |
|  | Mean | SD | Mean | SD | Mean | SD | Mean | SD |  |
| WBC | 5025 | 1047 | 7457 | 3237 | 11742 | 7205 | 14040 | 7806 | <0.001 |
| LYMPH | 1980 | 689 | 1782 | 1132 | 1432 | 1003 | 1088 | 1665 | <0.001 |
| MONO | 488 | 202 | 493 | 249 | 704 | 648 | 750 | 727 | 0.065 |
| Hb | 12.05 | 3.62 | 12.98 | 1.88 | 12.97 | 1.92 | 12.32 | 1.71 | <0.001 |
| Platelet | 208 | 39 | 256 | 92 | 256 | 105 | 290 | 158 | 0.732 |
| s.LDH | 208.25 | 12.23 | 219.39 | 45.66 | 293.47 | 164.27 | 488.70 | 279.33 | <0.001 |
| Index | 0.26 | 0.14 | 0.35 | 0.23 | 0.95 | 1.14 | 1.85 | 1.78 | <0.001 |

**Table 3:** Correlation between index and stage

|  |  | Index |  |  | P-value |
| --- | --- | --- | --- | --- | --- |
|  |  | Median | Mean | Standard Deviation |  |
| Stage | 1 | 0.259 | 0.26 | 0.14 | 0.0001 |
|  | 2 | 0.280 | 0.35 | 0.23 |  |
|  | 3 | 0.600 | 0.95 | 1.14 |  |
|  | 4 | 1.550 | 1.85 | 1.78 |  |
| Correlation Coefficient (r) = 0.41 |  |  |  |  |  |

**Figure 1:**
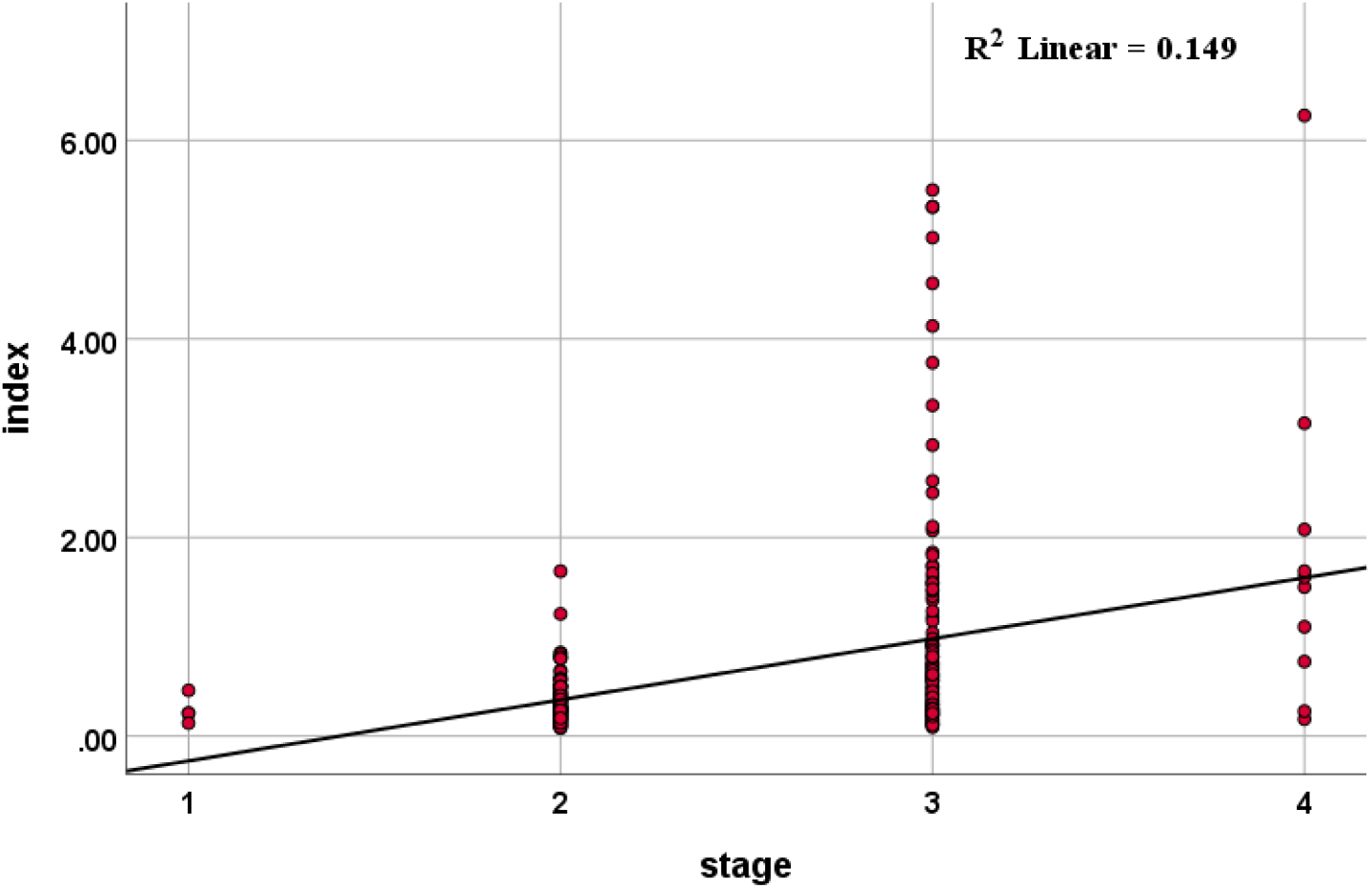
Dot plot represent the regression model between the index and the stage of Covid −19 disease.

## Discussion

We developed a new simple equation that depends on 3 main important parameters (serum lactate dehydrogenase, lymphocyte absolute count, and monocyte absolute count) to predict clinical outcome for patients with covid-19 (8,9), Our new index: COVID-19 severity-Iraqi-index (CSI).

LDH level serum improves the prediction power of Covid 19 severity. Also absolute lymphocyte count is good prognostic predictor of clinical outcome. Lymphopenia in patients with covid-19 can be explained by 2 mechanisms one of them is redistribution and the other is bone marrow selective lymphocyte suppression/

The new prognostic index of COVID-19 severity (CSI) has a very good correlation to the clinical stage. The higher the index the poorer the clinical outcome (p=<0.001).

We did not find similar index in the available published data regarding covid-19 prognosis. The score can be divided into low, intermediate, and high severity depending on the score number.

Any value for the index below 0.60 were considered as mild cases, while a score of 0.60 was considered as a cutoff point for moderate cases and a score of 1.55 as a cutoff point for severe cases.

This study has some limitations: some Other important parameters that were not included in our study are organ functions, CT scan results, and oxygen saturation because we intended to simplify the test early in the course of the disease. (10)

For instance Zhang et al pointed to the relation of CT chest involvement to many laboratory parameters like granulocyte count, LDH level, procalcitonin, and C reactive protein.(11)

A clinical correlation also mentioned in relation to progressive lung disease in relation to laboratory findings like lymphocytes, monocytes, platelets, and liver functions. (12) Others reported relation of clinical outcome and chest CT scan to D-dimer and C reactive protein.(13)

A positive relation also found between CT chest and leucocyte count, neutrophils and interlukin-2. (14, 15)

## Conclusions

COVID-19 severity-Iraqi index is a new simple predictor of clinical outcome in patients with covid-19 during early stage of the disease.

## Data Availability

Yes.We agree for availability of all data referred to in the manuscript.

## Contributors

All authors have contributed to the manuscript in study design, study concept, data collection and interpretation, and final writing and revision of the manuscript.

